# Plasminogenuria is associated with podocyte injury, edema, and kidney dysfunction in incident glomerular disease

**DOI:** 10.1101/19006809

**Authors:** Marc A. Egerman, Jenny S. Wong, Tian Runxia, Gohar Mosoyan, Kinsuk Chauhan, Fadi El Salem, Kristin Meliambro, Hong Li, Evren Azeloglu, Steven Coca, Kirk N. Campbell, Leopoldo Raij

**Affiliations:** Divison of Nephrology, Department of Medicine, Icahn School of Medicine at Mount Sinai; Katz Family Division of Nephrology and Hypertension, Department of Medicine, Univeristy of Miami Miller School of Medicine; Department of Microbiology, Biochemistry and Molecular Genetics, Rutgers University – New Jersey Medical School, Newark, NY, 07103, USA

**Keywords:** glomerular, podocytes, plasminogen, amiloride, edema, eGFR

## Abstract

Urinary plasminogen/plasmin, or plasmin(ogen)uria, has been demonstrated in proteinuric patients and exposure of cultured podocytes to plasminogen results in injury via oxidative stress pathways. A causative role for plasmin(ogen) as a “second hit” in kidney disease progression has yet to be demonstrated *in vivo*, and the association between plasmin(ogen)uria and kidney function in glomerular diseases remains unclear. We performed comparative studies in a puromycin aminonucleoside (PAN) nephropathy rat model treated with amiloride, an inhibitor of plasminogen activation, and measured changes in plasmin(ogen)uria and urinary endothelin-1 (ET1). In a glomerular disease biorepository cohort (n=128), we measured time-of-biopsy albuminuria, proteinuria, and plasmin(ogen)uria for correlations with renal outcomes. Increased glomerular plasmin(ogen) was found in PAN rats and FSGS patients. PAN nephropathy was associated with increases in plasmin(ogen)uria, proteinuria, and urinary ET1. Amiloride was protective against PAN-induced glomerular injury, reducing urinary ET1 and oxidative stress. In patients, we found associations between plasmin(ogen)uria and edema status as well as eGFR. Our study demonstrates a role for plasmin(ogen)-induced podocyte injury in the PAN nephropathy model, with amiloride having podocyte-protective properties. In one of largest glomerular disease cohorts to study plasminogen, we validated previous findings while suggesting a potentially novel relationship between plasmin(ogen)uria and eGFR. Together, these findings suggest a role for plasmin(ogen) in mediating glomerular injury and as a viable targetable biomarker for podocyte-sparing treatments.

**TRANSLATIONAL STATEMENT:** Proteinuria is associated with CKD progression, and increased cardiovascular morbidity and mortality. The underlying mechanisms of podocyte injury, the hallmark of proteinuric kidney disease, are poorly understood with limited, non-specific therapeutic options. This study adds to the evidence that plasmin(ogen) in the urine of proteinuric patients is associated with podocyte injury, edema, and impaired renal function. Previously published results from us and others, taken together with our current rodent model and human data, suggest that urinary plasmin(ogen) is a potential targetable biomarker. Efforts to decrease plasmin(ogen)-mediated podocyte injury could be part of a novel therapeutic strategy for glomerular disease.

## INTRODUCTION

Chronic kidney disease (CKD) affects approximately 500 million people worldwide, is increasing in prevalence, and disproportionately affects individuals from low- and middle-income countries.^1-3^ CKD significantly increases the risk of cardiovascular disease (CVD), which in turn is the leading cause of death in all stages of CKD.^4-7^ Independent of etiology, both proteinuria of increased severity and uncontrolled hypertension are associated with faster progression of CKD to end stage renal disease (ESRD).^8-12^ However, recent trials have shown that intensive blood pressure control, as compared to standard therapy, provides survival benefits associated with stroke and heart failure, but fails to prevent progression, particularly in proteinuric CKD patients with eGFR 30-45 ml/min/1.73m^2^ (Stage 3b) and beyond.^13^

Normal urine is essentially protein free due to the synergistic interactions at the glomerular filtration barrier between the fenestrated endothelium, the glomerular basement membrane, and the podocyte foot processes.^14-16^ Although playing an essential role in maintaining glomerular integrity, podocytes are highly susceptible to damage and have limited capacity for repair, with persistent proteinuria and progressive glomerulosclerosis occurring following excessive loss of the podocyte population.^17-22^ Despite significant evidence identifying podocytes as a key glomerular cell for injury, there remains a critical need for development of mechanistically-based strategies to promote podocyte survival and arrest CKD progression.^23-27^

Human and experimental rodent studies have demonstrated that persistent proteinuria is accompanied by abnormal excretion of urinary serine proteases of which plasminogen/plasmin is a principal component.^28-32^ Previous *in vitro* and *in vivo* reports have shown that the amiloride-sensitive sodium channel ENaC in the distal nephron is activated by plasmin, resulting in increased sodium reabsorption, thereby promoting volume expansion and hypertension.^33-37^ Studies in patients with CKD of diverse etiologies have confirmed a strong correlation between urinary plasminogen/plasmin levels—termed plasmin(ogen)uria—and sequela of volume overload, though most reports have concentrated on a limited number of subjects and/or diagnoses.^28,29,38-41^

Clinically it remains unclear whether progression of glomerular injury is driven strictly by the persistence of the initial insult and/or by a superimposed secondary pathogenic process, a so-called “second hit.” We recently proposed that filtered plasminogen may serve such a function, having demonstrated that cultured human podocytes express key mediators of the plasminogen-plasmin system—urokinase plasminogen activator (uPA), uPA receptor (uPAR), and tissue plasminogen activator (tPA)—and the novel plasminogen receptor PLG-RKT. More importantly, we showed that after binding, plasminogen is converted into plasmin through uPA, resulting in oxidative stress-mediated podocyte injury, directly via activation of NOX2/4 and indirectly through upregulation of endothelin-1 (ET1) synthesis.^42^ The latter is supported by previous studies linking ET1 to reciprocal endothelial/podocyte glomerular generation of reactive oxygen species (ROS) and the development of focal segmental glomerulosclerosis (FSGS).^43-46^ Of note, the ENaC inhibitor amiloride, with known off target effects inhibiting uPA-mediated plasminogen activation, was shown to arrest progression of plasmin(ogen)-mediated podocyte injury *in vitro*.^42,47,48^ Based on these findings, we hypothesized that persistent exposure to plasmin(ogen) serves as a “second hit” in glomerular disease and drive CKD progression, independent of the initial insult. In this manner, plasmin(ogen) may aggravate kidney disease both through direct injury to podocytes during its trans-glomerular passage and through hemodynamic stress linked to volume expansion via the sodium retention effects of ENaC in later stage disease.

To this end, we tested whether our initial *in vitro* results from cultured podocytes could be translated experimentally to puromycin aminonucleoside (PAN) nephropathy, a well-established *in vivo* model of podocyte injury, and clinically by investigating cross-sectional associations between plasmin(ogen)uria and renal disease characterics in one of the largest cohorts of glomerular diseases to study plasmin(ogen). Here, we (i) present strong supportive evidence for a causative role of the plasmin(ogen)-system in podocyte injury, with amiloride having reno-protective properties *in vivo*, and (ii) advance clinical correlations of plasmin(ogen)uria as a biomarker of glomerular injury in proteinuric patients. Importantly, given such a function, the plasmin(ogen) pathway represents an attractive target for the development of mechanistically-based novel therapeutic interventions.

## RESULTS

### Amiloride decreases proteinuria and urinary endothelin in PAN nephropathy

The PAN nephropathy model of podocyte injury has been previously shown to be associated with increased urinary plasmin(ogen).^35^ To reproduce these findings and determine the effects of amiloride treatment, adult male Wistar rats (6-8 weeks old) were treated with a single intravenous dose of PAN (100 mg/kg), PAN plus amiloride (0.5 mmol/L), or vehicle, and sacrificed on day eight. As expected, PAN treated rats developed significant proteinuria compared to controls (6.84±0.62 mg/mg·Cr vs. 0.68±0.07 mg/mg·Cr; p<0.0001). Treatment with amiloride was associated with reduced proteinuria, as compared to PAN alone (3.11±0.32 mg/mg·Cr; p<0.0001) (Figure 1A). In addition, PAN treatment caused increases in both plasmin(ogen)uria (15.96±2.47 µg/mg·Cr vs. 2.16±0.49 µg/mg·Cr; p<0.0001) (Figure 1B) as well as urinary ET1 (330.5±30.6 pg/mg·Cr vs. 3.05±0.28 pg/mg·Cr; p<0.0001) (Figure 1C). Of note, amiloride reduced PAN-induced plasmin(ogen)uria by 22.0%, but the difference was not statistically significant (p=0.095).. However, amiloride was associated with a significant reduction in urinary ET1 excretion (330.5±30.6 pg/mg·Cr vs. PAN alone: 30.37±2.81 µg/mg·Cr; p=0.175; p<0.0001) (Figure 1B-C). Thus, PAN nephropathy is associated with increased urinary plasmin(ogen) and ET1, the latter of which was rescued by amiloride.

**Figure 1.**
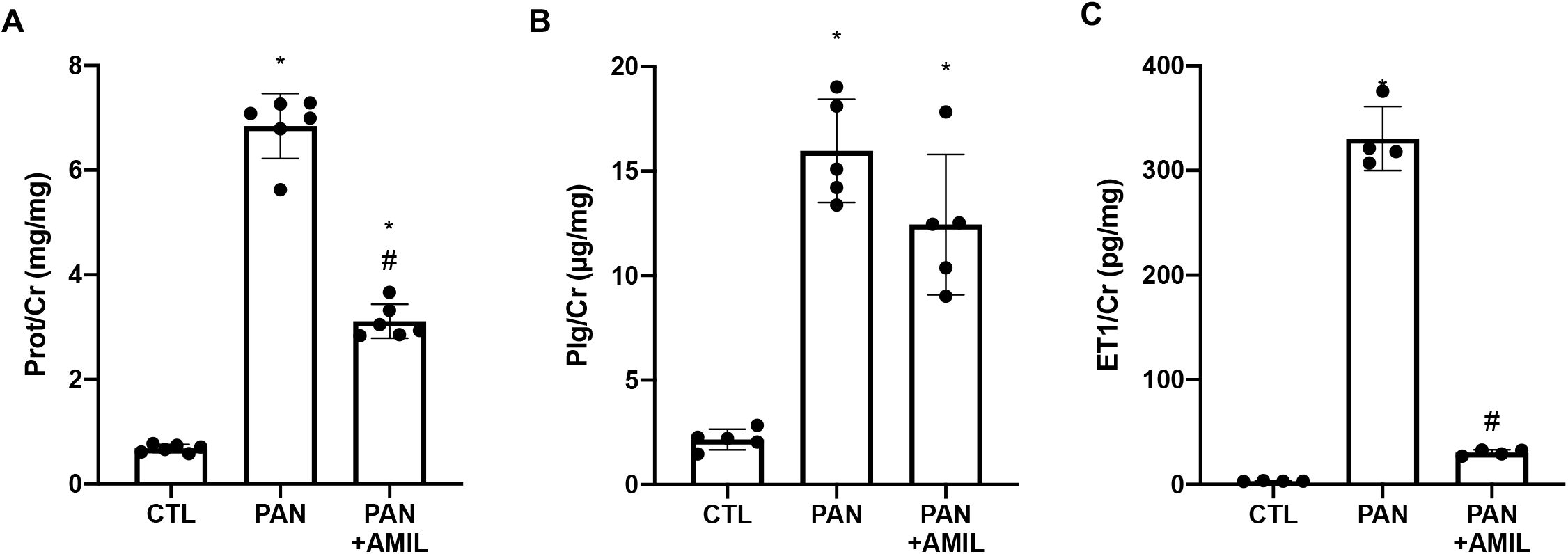
PAN nephropathy promotes urinary excretion of total protein, plasmin(ogen), and endothelin-1. PAN caused increased total proteinuria (**a**), plasmin(ogen)uria (**b**), and urinary ET1 (**c**). Administration of amiloride (0.5mmol/L amiloride q.d.) significantly reduced total protein (**a**) and endothelin excretion (**c**). Plg/Cr, urinary plasmin(ogen)/creatinine; ET1/Cr, urinary endothelin/creatinine; Prot/Cr, urinary protein/creatinine; amil, amiloride; * p<0.05 relative to CTL; # p< 0.05 relative to PAN alone

### Glomerular endothelin-1 and plasmin(ogen) are increased in PAN nephropathy

Both endothelin-1 and its receptors ETA/ETB have been identified within the glomerulus, where ET1 exerts a pro-injurious effect.^45,49,50^ We recently demonstrated that ET1 can act under the influence of plasmin(ogen) in an autocrine/paracrine fashion to induce oxidative stress in cultured podocytes.^42^ Given the reversibility of PAN nephropathy induced by a single injection,^51-53^ a separate time course experiment was conducted in order to objectively trend the time-dependent nature of glomerular plasmin(ogen). Across two proteomic analyses, the average size of the top 50 enriched proteins by mass-spectrometry were 85±56 kDa and 59±40 kDa, respectively, suggesting a wide range of identified molecular weight proteins. High plasmin(ogen) levels were identified in glomeruli of PAN-treated rats—glomerular plasminogen was elevated on day 3 and peaked on day 7 before returning to baseline by day 14-21 (Figure 2A). The peptides identified as plasminogen were from both the N- and C-terminal regions (data not shown). We confirmed the presence of significant levels of plasmin(ogen), in addition to increased glomerular expression of ET1, within glomerular isolates by western blotting (Figure 2B). Taken together, excessive filtration of plasminogen occurs early following PAN-mediated podocyte injury, whereupon plasmin(ogen) strongly binds within the glomerulus.

**Figure 2.**
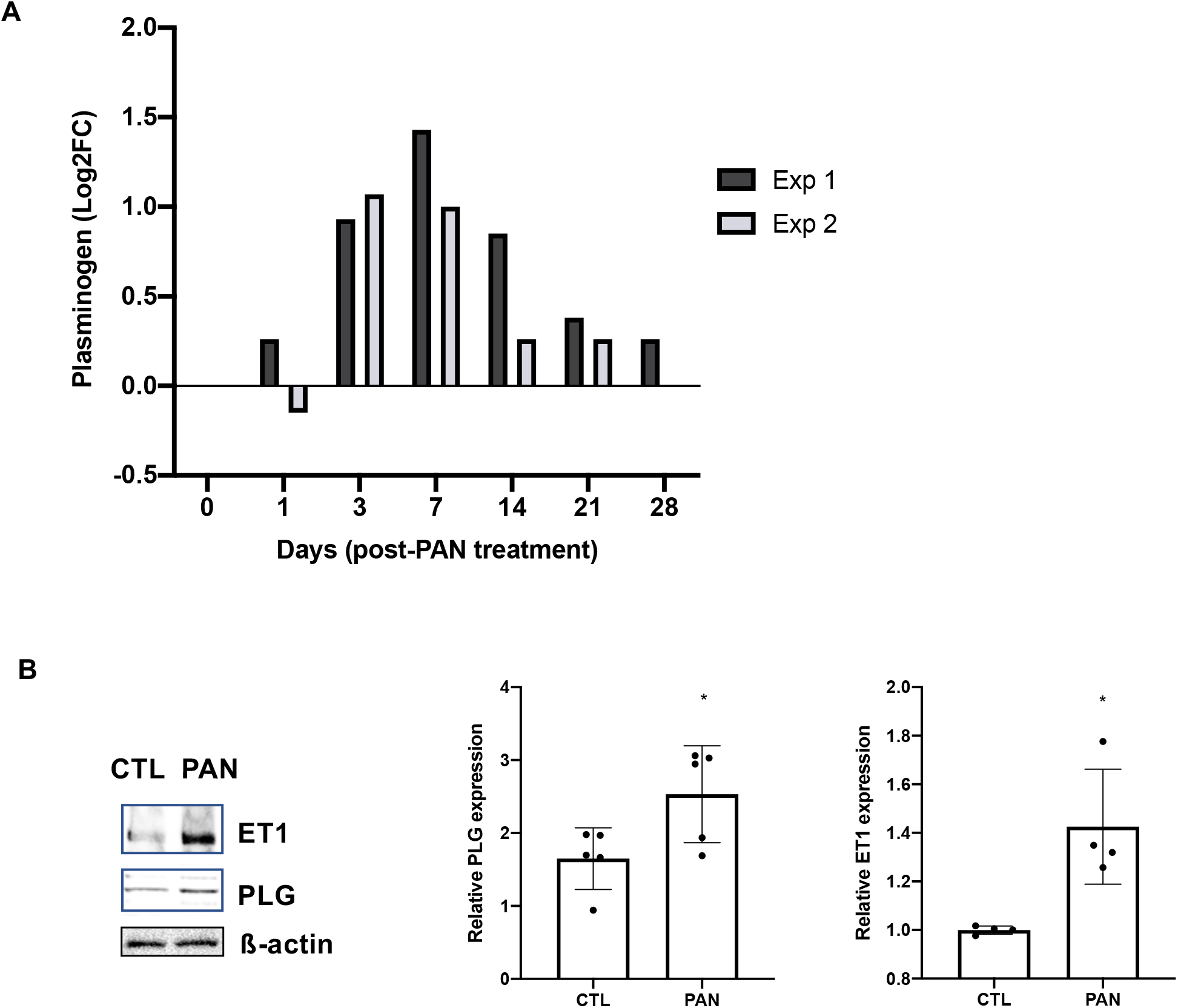
PAN nephropathy increases glomerular plasmin(ogen) and ET1. Glomeruli from PAN treated rats were isolated for mass spectrometry or immunoblotting. (**a**) Plasmin(ogen) levels increased early following single dose PAN treatment by proteomics; dark gray box, experiment 1; light gray box, experiment 2. Plasmin(ogen) and endothelin-1 levels were increased in glomeruli of PAN-treated rats as detected by western blotting (**b**). Representative images are shown. The abundance of β-actin was used to normalize and densitometric analysis was performed using ImageJ. Each bar represents the mean ± SD across 4-5 rats. PLG, plasmin(ogen); ET1, endothelin-1; * p <0.05

### Amiloride treatment decreases oxidative stress and improves podocyte integrity in PAN nephropathy

We previously demonstrated that plasmin(ogen)-mediated ROS generation involved podocyte-specific upregulation of NADPH oxidases NOX2/4 and the B scavenger receptor CD36.^42^ Here, we sought to determine the oxidative stress changes associated with PAN treatment with or without amiloride. By immunofluorescence staining, we found that PAN treatment increased glomerular CD36, which colocalized with the podocyte marker synaptopodin (Figure 3A). Glomerular gene expression by qPCR also showed increased CD36 with PAN (data not shown). Similarly, we confirmed PAN-mediated ROS generation in podocytes with enhanced double labeling immunofluorescence for 8-OxoG and synaptopodin (Figure 3B). PAN-induced increases in CD36 and 8-oxo-G levels were significantly ameliorated by amiloride treatment (Figures 3A-B).

**Figure 3.**
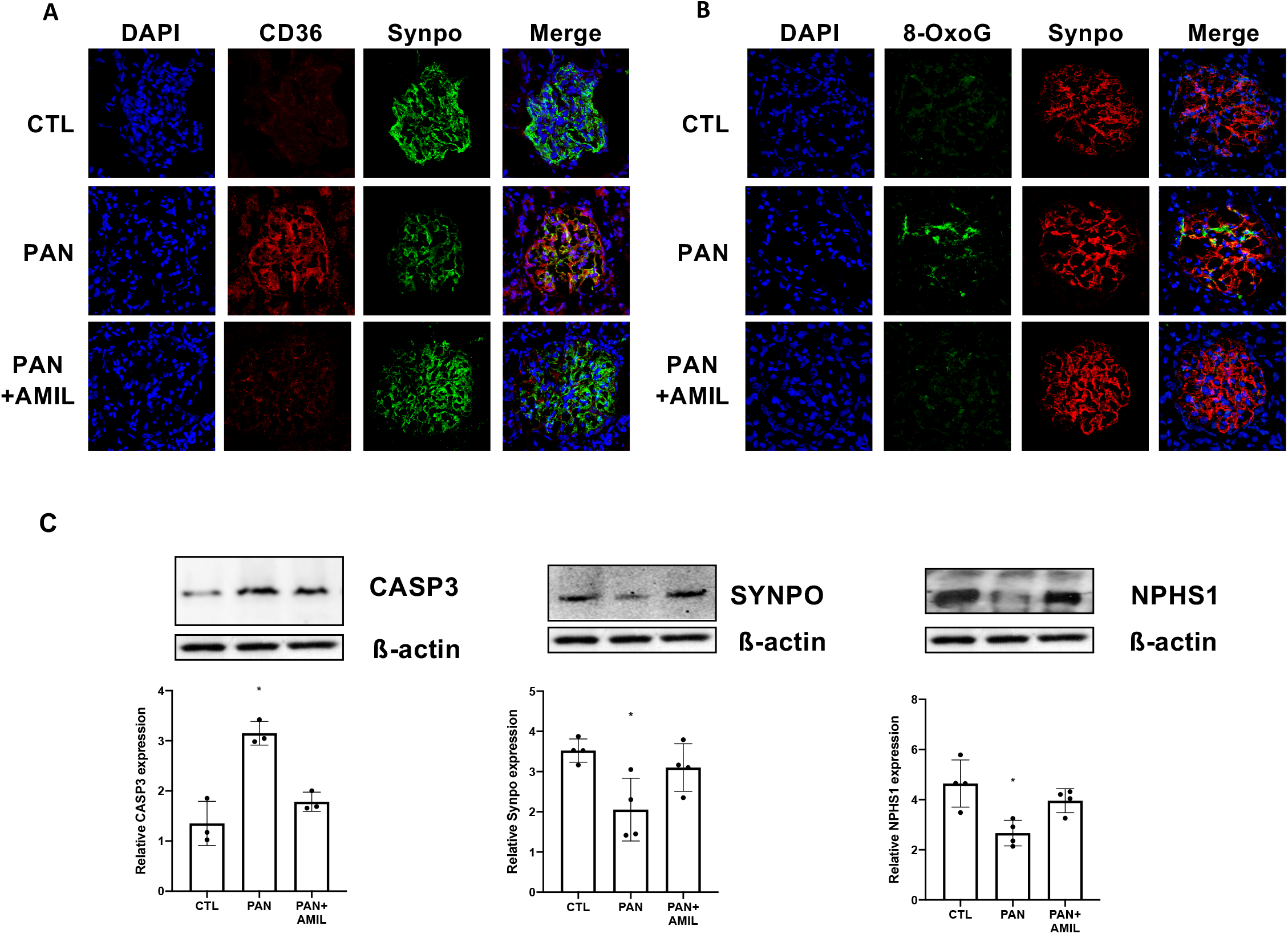
PAN nephropathy is associated with increased oxidative stress and impaired podocyte homoeostasis. (A, B) CD36 and 8-OxoG robustly accumulated in podocytes following PAN treatment. Administration of amiloride caused dramatic reduction in both markers. Red color shows staining for CD36 (A) and the podocyte marker synaptopodin (B). Green color shows staining for synaptopodin (A) and 8-Oxo-G (B). Nuclei were colored blue with DAPI. (C) Significant bands for caspase 3, synaptopodin, and nephrin were observed in samples of isolated glomeruli from PAN-treated rats. Representative images are shown. The abundance of β-actin was used to normalize and densitometric analysis was performed using ImageJ. Each bar represents the mean ± SD across 3-4 rats. * p<0.05

Studies by us and others have highlighted the contribution of podocyte apoptosis to glomerular disease progression.^21,22,42^ In support of our previous study, which showed plasminogen-mediated podocyte apoptosis augmented by oxLDL, we found that total glomerular caspase 3 expression was enhanced with PAN treatment and reduced with amiloride (Figure 3C). Moreover, we determined that glomerular levels of other markers of podocyte integrity, specifically nephrin and synaptopodin, were reduced in PAN nephropathy (Figure 3C). Of note, both nephrin and synaptopodin are sensitive to ROS mediated injury.^54,55^ Overall, the pathophysiologic changes observed with PAN treatment were either abrogated or reduced with amiloride (Figure 3). Taken together, these findings suggest that amiloride is at least partly protective/stabilizing against PAN-induced oxidative stress and glomerular injury.

### Urinary plasmin(ogen) excretion is increased in human glomerular disease

Our study cohort included 128 patients with glomerular diseases with urine samples collected at the time of kidney biopsy. The full characteristics of the cohort are summarized in Table 1. Subjects had an average age of 48.3 years with mean eGFR 57.5 ± 36.7 ml/min/1.73 m^2^. Of the subjects, 27% were edematous with 32% and 47% prescribed diuretics and ACEi/ARB, respectively, at the time of biopsy. Participants with urinary plasmin(ogen) (log µg/mg•Cr) in the upper quantile tended to be older with lower eGFR and were more likely to have edema with a prescription for diuretics and ACEi/ARB (Table 1).

**Table 1.**
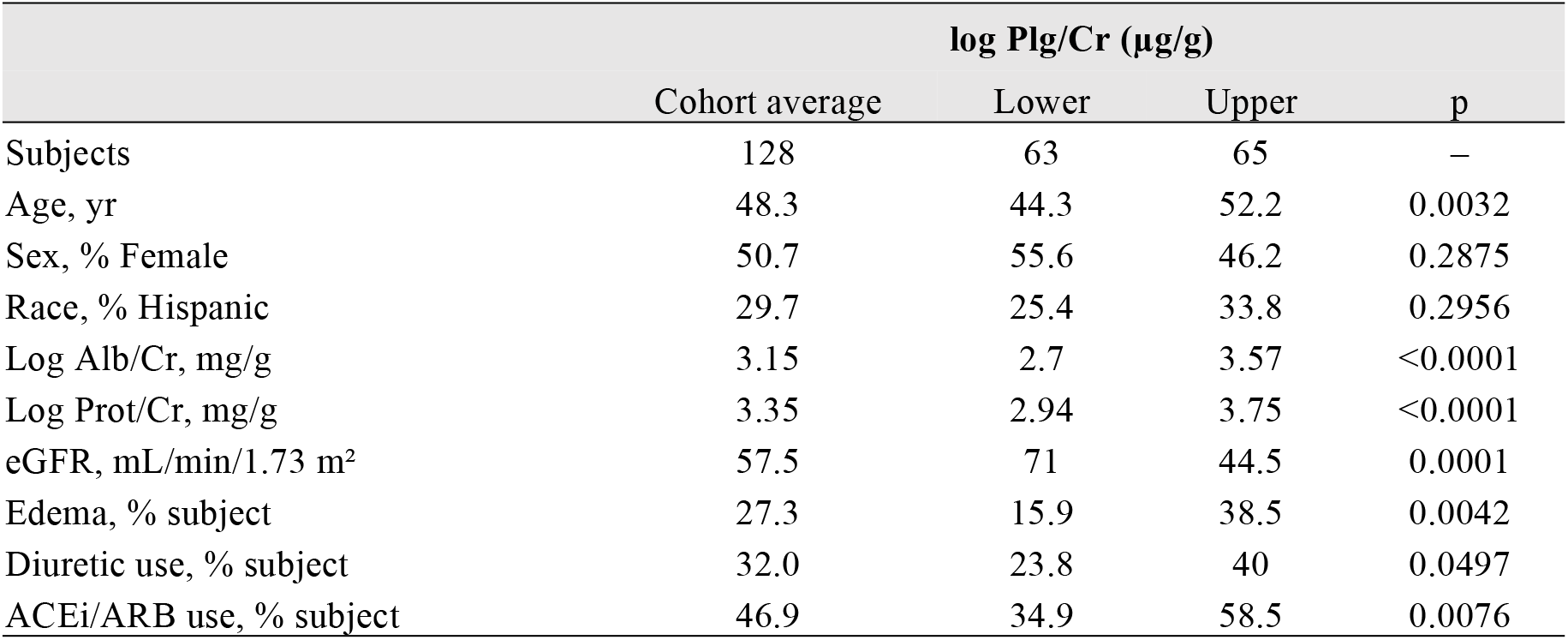
Study characteristics of glomerular disease patient cohort. Values reported are n (percentages) for categorical variables and means for continuous variables. Data are presented as means stratified on the basis of uPl/Cr quantiles. eGFR is the GFR determined using the CKD-EPI formula. Alb/Cr, urinary albumin/creatinine; Prot/Cr, urinary protein/creatinine; Plg/Cr, urine plasmin(ogen)/creatinine

Urinary plasmin(ogen) has been previously shown to be associated with hypervolemia in limited patient cohorts.^28,29,38-41^ To validate these findings across a larger sample size in the setting of biopsy-proven glomerular diseases, we sought to define these relationships within our cohort. At baseline across all subjects, plasmin(ogen)uria (log µg/mg•Cr) was strongly positively correlated with both proteinuria (log mg/mg•Cr; r=0.8348, p<0.0001) and albuminuria (log mg/mg•Cr; r=0.8219, p<0.0001) (Figure 4). In addition, plasmin(ogen)uria had an inverse correlation with eGFR (r=-0.3320, p=0.0001) (Figure 4). To examine whether relative urinary plasmin(ogen) was associated with edema status and kidney function, we compared plasmin(ogen)uria with cross-sectionally collected data. Higher levels of all three biomarkers— plasmin(ogen)uria, albuminuria, and proteinuria—were associated with the presence of edema. However, only plasmin(ogen)uria was significantly lower in patients with eGFR ≥ 60 ml/min/1.73 m^2^, compared those with eGFR < 60 ml/min/1.73 m^2^ (Figure 4).

**Figure 4.**
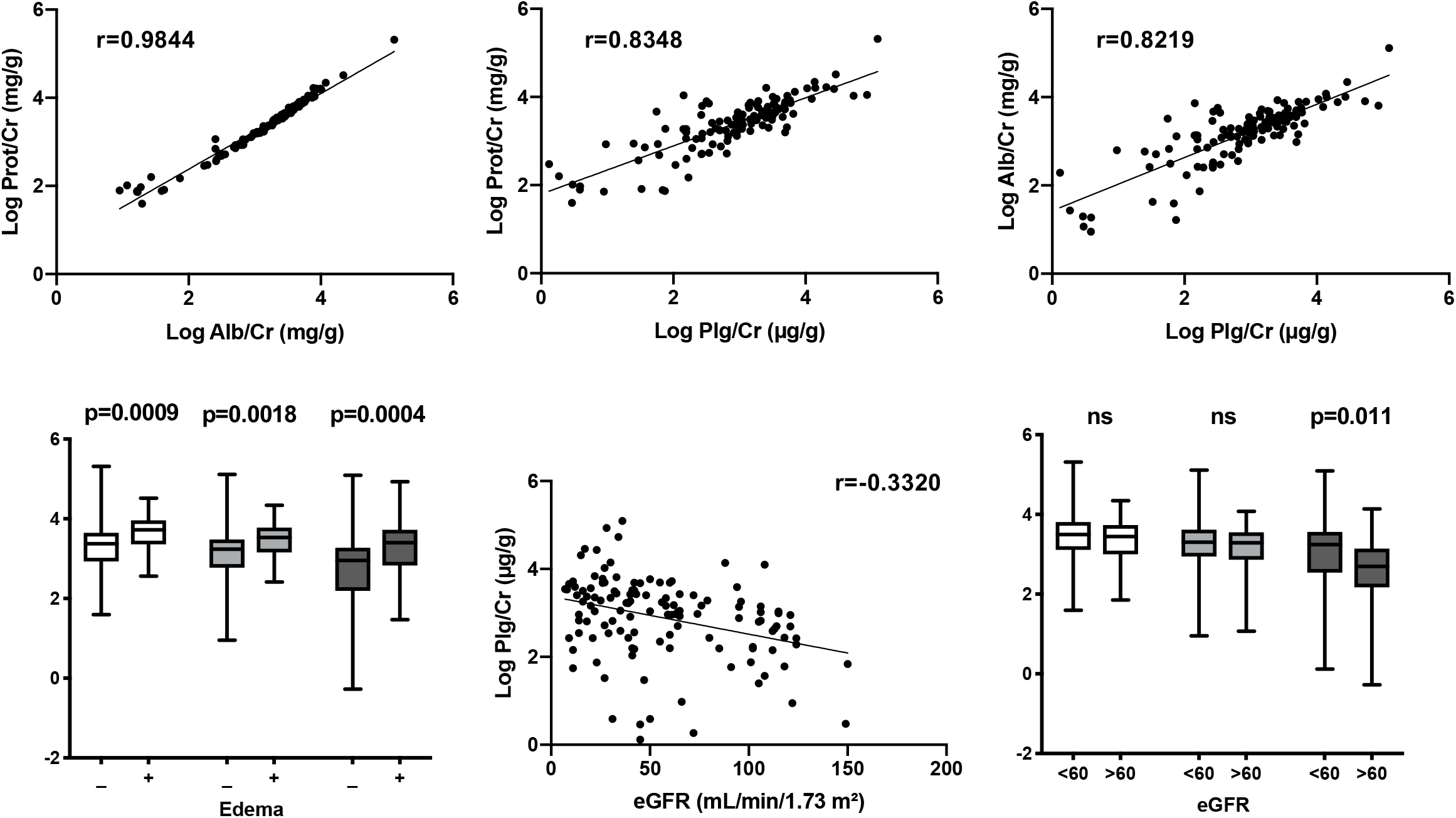
Plasmin(ogen)uria correlates with both urinary biomarkers and kidney disease outcomes in cross-sectional analysis of patient urines. Log of urinary plasmin(ogen)/creatinine (log Plg/Cr) correlated with both total urine protein/creatinine (log Prot/Cr) and albumin/creatinine (log Alb/Cr) at biospy. The presence of edema was significantly associated with higher levels of all tested urine markers whereas high log Plgl/Cr correlated weakly with lower eGFR at time of biopsy. The ordinate represents log units, but the units for log Plg/Cr (μg/g) differ from those of log Prot/Cr and log Alb/Cr (mg/g). Whiskers represent standard deviations. White boxes, log Prot/Cr; light gray boxes, log Alb/Cr; dark gray boxes, log Plg/Cr

In univariate analysis, plasmin(ogen)uria, along with both albuminuria and proteinuria, were similarly associated with increased odds of edema at time of biopsy (Table 2). Adjusting for age, gender, race, use of ACEi/ARB or diuretics, and baseline eGFR did not significantly affect the observed strengths of association. The three biomarkers had similar negative correlations with eGFR at time of biopsy, all of which were modestly attenuated after adjustment for covariates. (Table 2).

**Table 2.** Urine biomarkers are associated with edema and eGFR at time of biopsy. (a) All three markers tested (log Prot/Cr, log Alb/Cr, log Plg/Cr) correlated with increased odds of edema at biopsy in both univariate and adjusted regression models. (b) All three markers tested (log Prot/Cr, log Alb/Cr, log Plg/Cr) were associated with reduced eGFR in both univariate and adjusted regression models. ^a^Variables selected for adjustment by *a priori* specification; included age, gender, race/ethnicity, use of drugs of time of biopsy: ACEi/ARB, diuretics, and eGFR (for edema model)

| Predictor | Edema OR (95% CI); p value |  |
| --- | --- | --- |
|  | Univariate | Multivariable<sup>a</sup> |
| Proteinuria, log mg/g Cr | 3.81 (1.74 to 9.73); p=0.0022 | 3.80 (1.58 to 10.66); p=0.0056 |
| Albuminuria, log mg/g Cr | 3.43 (1.61 to 8.75); p=0.0041 | 3.49 (1.50 to 9.66); p=0.0079 |
| Urinary plasmin(ogen), log µg/g Cr | 2.58 (1.53 to 4.79); p=0.0010 | 2.96 (1.56 to 6.35); p=0.0023 |

**Table 2 | Urine biomarkers are associated with edema and eGFR at time of biopsy.**
| Predictor | eGFR (95% CI); p value |  |
| --- | --- | --- |
|  | Univariate | Multivariable<sup>a</sup> |
| Proteinuria, log mg/g Cr | -14.78 (-24.63 to -4.93); p=0.0036 | -8.11 (-17.40 to 1.17); ns (p=0.09) |
| Albuminuria, log mg/g Cr | -11.04 (-19.83 to -2.24); p=0.014 | -6.10 (-14.29 to 2.09); ns (p=0.14) |
| Urinary plasmin(ogen), log µg/g Cr | -13.82 (-19.76 to -7.88); p<0.0001 | -9.74 (-15.36 to -4.122); p=0.0008 |

### Glomerular plasmin(ogen) is increased in human FSGS

Given our results demonstrating glomerular plasmin(ogen) in the PAN nephropathy rodent model, we sought to initially translate this novel finding to the clinical setting. Consistent with our observations in PAN-treated rats, we observed significantly more plasmin(ogen) within glomeruli from a series of human kidney biopsies from patients with FSGS, as compared to age-matched normal control kidney tissue (Figure 5). Thus, as in rodents, plasmin(ogen) is strongly associated with the glomerulus following damage to the glomerular filtration barrier.

**Figure 5.**
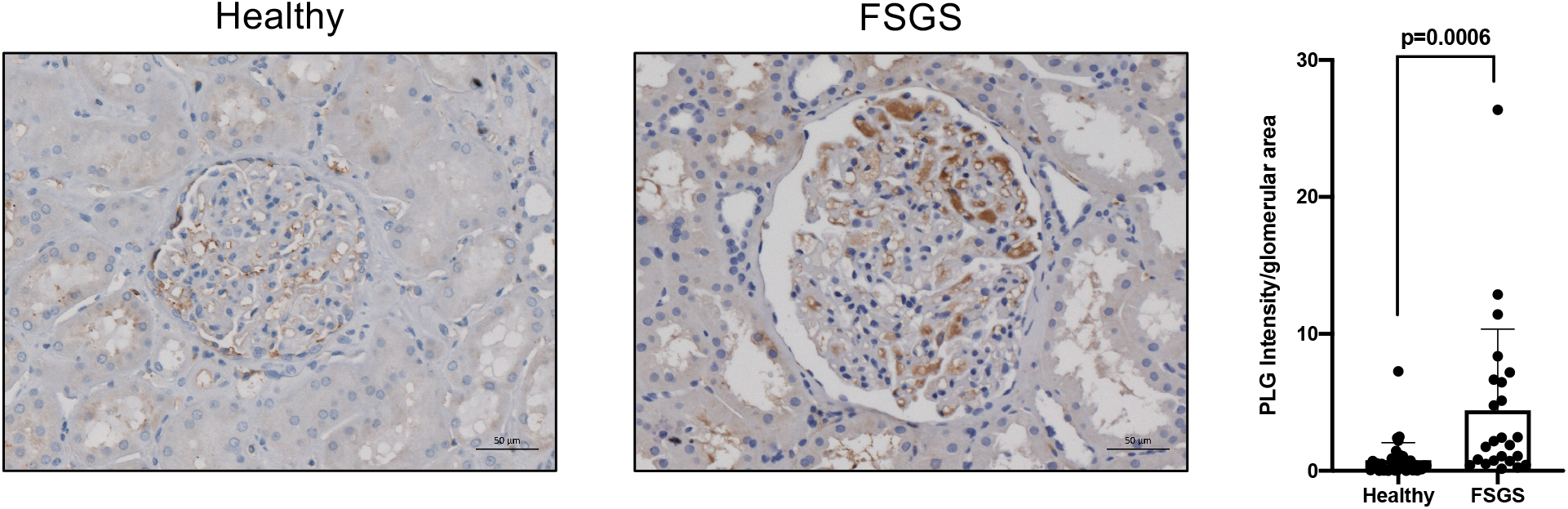
Immunohistochemistry of kidney from patients with focal segmental glomerulosclerosis (FSGS) is consistent with glomerular-associated plasmin(ogen) (a) Immunostaining for plasmin(ogen) was performed on paraffin-embedded human kidney sections. Representative images are shown. Bar = 50 μm. (b) Quantification of glomerular staining intensity was calculated and is shown as a fold change relative to control samples. Glomerular plasmin(ogen) staining is significantly increased in patients with FSGS compared with control subjects. Total number of glomeruli analyzed: n = 37 control (9-14 glomeruli from each of 3 individual patients), n = 24 FSGS (4-10 glomeruli from each of 3 individual patients); scale bar = 50 µm

## DISCUSSION

In the present study, we (i) experimentally assessed whether our previously published *in vitro* results in human podocytes regarding plasminogen and amiloride, the inhibitor of its conversion/activation, could be translated to a well-established *in vivo* proteinuric rodent model and (ii) sought to better define the associations between plasmin(ogen)uria and clinically relevant kidney disease characterics in patients with glomerular diseases. Collectively, our findings, together with previous *in vitro* data, lends further support to a model in which urinary plasmin(ogen) acts a “second hit” with direct and/or indirect effects on glomerular integrity in promoting CKD progression.

Extensive studies in animal models, with supporting evidence from nephrotic patients, suggest that, in the setting of proteinuria, plasma-derived plasminogen is aberrantly filtered and converted to plasmin by uPA, thereby activating the amiloride-sensitive sodium channel ENaC in the distal nephron to enhance renal sodium retention, independent of the renin-angiotensin-aldosterone system.^28-32^ The current paradigm for urinary plasminogen has thus primarily focused for the most part on its role in the collecting ducts and its contribution to volume overload status, in large part due to limited studies exploring the potential role of plasmin(ogen)-induced podocyte injury in proteinuric glomerular injury.^38-41,56,57^

It is well established that independent of baseline eGFR or etiology, total proteinuria is a marker of kidney damage and a strong predictor of progression to end‐stage renal disease, with increasing proteinuria in the setting of CKD correlating with increased risk of cardiovascular disease and all-cause mortality.^58-63^ Few reports, however, have identified a role for specific filtered proteins that contribute to kidney damage through cytotoxic effects on podocytes as “second hits.” Such studies, however, include the identification of albumin as a potential contributor of both glomerulosclerosis and tubulointerstitial injury via direct and indirect mechanisms.^64-66^

In support of our previously published data in cultured human podocytes, which suggested that filtered plasminogen may in fact serve such a role,^42^ we induced PAN nephrosis in rats, a model in which ROS-mediated cytoskeletal dysfunction has been documented to promote foot process damage.^42,51,67,68^ Mechanistically, we found that PAN treatment increased markers of glomerular oxidative stress and apoptosis, likely due at least in part to alteration in NOX2/4 activity based upon our previous results.^42^ PAN treatment was also associated with a concurrent reduction of glomerular levels of synaptopodin and nephrin, two critical regulators of podocyte functional and structural homeostasis. Consistent with prior clinical and experimental studies linking ET1 to glomerular ROS generation,^45,46,49^ we detected ET1 expression in glomeruli isolates following PAN treatment. Additionally, we determined—for the first time to our knowledge—that there was significant plasmin(ogen) within glomeruli of PAN rats and FSGS patients, strongly suggesting that plasmin(ogen) is capable of binding and therefore mediating pro-injurious effects *in situ* on the glomerular filtration barrier. These findings are in support of the ability of plasminogen to bind podocytes, given our observed recapitulation of downstream effects from *in vitro* models, specifically in terms of increased markers of oxidative stress and decreased markers of podocyte homeostasis following PAN treatment.^42^

PAN-treated rats developed severe proteinuria as expected, in addition to significant plasmin(ogen)uria and urinary excretion of ET1. This is in conjunction with previous reports of high levels of urinary plasmin(ogen) following PAN administration and similar to rodent models of FSGS and HIV-associated models of nephropathy.^35,42^ Following amiloride treatment, we observed significant declines in both total proteinuria and urinary ET1, with a trend toward a decrease in plasmin(ogen)uria. The observed plasmin(ogen)uria results are likely due to the inability of the immunoassay to discriminate between plasminogen and plasmin, given the effect of amiloride in changing the relative plasmin/plasminogen ratio, as opposed to total plasmin(ogen) levels.^35^

The rescue phenotype observed with amiloride was consistently observed throughout our study. Previous reports on amiloride initially demonstrated that it potently inhibits urinary uPA activity, thereby preventing plasmin-mediated stimulation of ENaC activity and sodium retention.^29,31,35^ Our data demonstrates that amiloride may have direct podocyte-sparing properties *in vivo*, upstream of its ENaC functionality, given that the only published report linking an anti-oxidant effect with amiloride is dependent on its regulation of plasmin(ogen)-mediated oxidative stress.^42^

In a model of HIV induced podocytopathy-associated proteinuria, unilateral ureteral obstruction was induced for one week, which mechanically arrested GFR as well as trans-glomerular passage of proteins, including plasminogen. This maneuver resulted in ipsilateral preservation of podocyte morphology, but contralateral progressive podocytopathy, glomerulosclerosis, and proteinuria.^69^ Given that high levels of plasmin(ogen)uria have been observed in the HIVAN model, this study lends further support to the hypothesis of plasmin(ogen) as a “second hit,” wherein mechanical prevention of podocyte exposure to aberrant proteinuria and plasmin(ogen)uria provides podocyte/glomerular protection.

In addition to serving as a putative causative agent in the glomerular disease process, studies in patients with CKD of various etiologies have both detected the presence of excessive urinary plasmin(ogen) and demonstrated its role as a novel biomarker.^28,29,38-41^ Plasmin(ogen)uria has been correlated with markers of glomerular proteinuria, namely albuminuria, and found to be associated with sequela of fluid overload, including hypervolemia and hypertension, though most studies have been limited by sample size and/or single-disease etiologies.^38-40,70^

In our cohort of 128 patients with biopsy-proven podocytopathies, we examined the association between urinary plasmin(ogen) levels and both volume overload (as determined by edema status) and kidney function (as determined by eGFR), compared with both albuminuria and proteinuria. The correlation we observed between plasmin(ogen)uria and albuminuria (r=0.8219) is consistent with previous reports.^12,23^ We also observed that urinary excretion of plasmin(ogen) was independently correlated with edema status at time of biopsy, as are both total proteinuria and albuminuria, with all urine markers exhibiting similar degrees of association. These cross-sectional results are similar to previous findings in the later course of disease, including in patients with stable CKD.^29,39,40^ We also observed inverse relationships between the three biomarkers and eGFR at biopsy. This represents the first time—to our knowledge—of a potential relationship between urinary plasmin(ogen) excretion and eGFR in the setting of biopsy-proven glomerular diseases. Although the cross-sectional nature of our study prevents conclusions as to a direct contribution of plasmin(ogen) to changes in eGFR, such a relationship is of interest, given the likely pro-injurious role of plasmin(ogen) on the glomerulus. Importantly, plasmin(ogen)uria behaves similar to the other urine markers tested, relative to its associations with clinical characteristics, but it remains the most practical to be targeted by therapeutic intervention, as compared to urinary albumin.

The results of this study should be viewed in light of some limitations. Our rodent model is associated with significant plasmin(ogen)uria, with amiloride having rescue effects that were previously attributed to plasmin(ogen) *in vitro*.^42^ Although other proteins and/or mechanisms may account for some of the observed results, our data provides indirect evidence for a role of plasmin(ogen) in mediating injury in the PAN model. We were unable to identify relative changes in plasminogen vs. plasmin given the immunoassay employed—prior studies have used western blotting analysis for differentiation between plasminogen and plasmin, which was not a feasible for a large number of samples in the current study.^29,35^ Because of collinearity, proteinuria, albuminuria, and plasmin(ogen)uria could not be included in the same models and, as such, independent effects could not be properly assessed statistically. However, the proposed biological mechanism supports a direct role plasmin(ogen). In addition, the goal of study was not necessarily to find an independent biomarker, but rather a biomarker with likely underlying causative role in the disease process. Our cohort is limited in size and represents only cross-sectional data, thereby preventing more sophisticated sup-group analyses and time-to-event outcomes. However, it remains one of the largest to study plasmin(ogen) to date across biopsy-proven etiologies and lays the foundation to study the relationship between plasmin(ogen)uria and eGFR longitudinally in a similar population.

Overall, our animal data suggest a possible role of plasmin(ogen)uria in mediating kidney injury, at least in part through oxidative stress pathways. In combination with our clinical findings and previous reports, this suggest a dual role of plasmin(ogen) in the progression of kidney disease as a targetable biomarker, with effects at two independent sites within the nephron. As such, our results may signal a shift in the current paradigm from the view of targeting plasmin(ogen)uria to solely treat sodium retention in nephrotic syndrome to early inhibition of plasmin activity in promote glomerular integrity in a wide-range of podocytopathies. Amiloride may reduce proteinuria only modestly, but offer protection from progressive injury to surviving podocytes, provided by the critical inhibition of uPA and proteolytic activation of plasminogen to plasmin. Dose-finding studies to determine whether lower concentrations of amiloride provides glomerular protective effects with reduced risks of complications attributed to doses used in the setting of resistant edema may be appropriate.

## METHODS AND MATERIALS

### Animal Studies

PAN nephropathy was induced as per established protocol.^71,72^ Briefly, 6-8 weeks old male Wistar rats were intravenously injected either with either PAN (n=7; single IV 100 mg/kg) or PAN + amiloride (n=8; PAN + 0.5mmol/L amiloride). PBS-injected age-matched rats (n=6) served as controls. Animals were euthanized 7-8 days after PAN injection. Kidneys were removed and glomeruli were isolated by serial sieving in ice-cold PBS. All animal protocols were approved by IACUC at the Icahn School of Medicine at Mount Sinai and the Miller School of Medicine at University of Miami.

### Study participants

Subjects with biopsy-proven diagnoses (diabetic, IgA, membranous, nephropathy; lupus nephritis, FSGS, arterionephrosclerosis) were recruited and provided written informed consent at enrollment, as per the approved Institutional Review Board (IRB). Freshly voided urine samples were collected, spun at 3000 rpm for 15 minutes. Supernatants were aliquoted and stored at −80°C prior to use. All clinical data was collected from electronic medical records. Frozen sections from archived human biopsy material were obtained from the Icahn School of Medicine at Mount Sinai under IRB approved protocol. All biopsies were clinically indicated, and only extra tissue not required for diagnostic purposes was permitted for research use; therefore, no patient-informed consent was required.

### Urinary Biomarker Determination

Rat urine endothelin and plasmin(ogen) were measured using commercial kits (Cayman, Item No:583151; ICLLAB, E-25PMG, respectively), as per protocol. Rat urine creatinine was measured according to the manufacturer’s protocol (Cayman, Item No: 500701). For human samples, plasmin(ogen) from urine at time of biopsy was measured using a commercial kit (Innovative Research, IHPLGKT-TOT), as per protocol. Concentrations were estimated from a standard curve and normalized by urine creatinine. All other patient values including creatinine were determined by the chemistry laboratory at Icahn School of Medicine at Mount Sinai.

### Western blotting, IF, IHC

For westerns, samples were processed by standard protocol. Briefly, tissue was homogenized and protein quantified by Bradford assay. Equal amounts of protein were loaded and separated by SDS-PAGE and transferred to nitrocellulose. Data were normalized by β-actin and expressed as fold-change relative to control. Anti-synpo was a gift from Dr. Peter Mundel (MGH), all other primary antibodies were purchased from Santa Cruz. For immunostaining, sections were cut on slides which were blocked and incubated with the appropriate primary and secondary antibodies, as per standard protocol. Secondary Alexa conjugated antibodies were purchased from Thermo Fisher (Life Technologies). Nuclei were labeled with DAPI. All primary antibodies for IF were purchased from Santa Cruz. Standard DAB IHC was performed at Mount Sinai Department of Pathology, using Ventana Discovery Ultra IHC/ISH research platform. Antigen retrieval was achieved with proprietary CC1 solution (Roche Diagnostics) for 1 hr. Primary antibody against plasmin(ogen), which was diluted in PBS 1:400, was manually applied and incubated for 1 hr, followed by secondary anti-rabbit antibody (Roche). Counterstain was DAB.

### Proteomics

Isobaric labelled shotgun proteomics was performed as previously described.^71^ Briefly, samples were lysed in urea with protease and phosphatase inhibitors and proteins were tryptically digested overnight. Peptides were labeled using iTRAQ isobaric tags (Thermo Scientific) per manufacturer’s instructions and were desalted and fractionated. Liquid chromatography with tandem mass spectrometry (LC-MS/MS) was performed using an UltiMate 3000 LC System and LTQ Orbitrap Velos mass spectrometer. MS/MS spectra were analyzed using Mascot and Sequest search engines with the Proteome Discoverer platform. A false discovery rate cutoff of 0.01 was used to limit the forward/reverse database searches that yielded peptide identification with a 95% confidence interval. Relative quantification was performed on the Scaffold platform using the log2 values of iTRAQ label ratios.

### Clinical Outcomes and Statistical Analysis

Cross-sectional primary outcomes were edema status or eGFR at time of biopsy. Correlations between biomarkers and relevant characteristics were assessed using Spearman’s partial correlations. The association of each biomarker with the kidney-related clinical characteristic was evaluated (log base 10–transformed) by univariate and multivariable logistic and linear regression methods (adjusted for age, gender, race/ethnicity, RASi or diuretic use, and eGFR, as appropriate)

## Data Availability

Raw data available upon request

## ACKNOWLEDGEMENTS

We thank Siarhei Szedzik for immunohistochemistry work. We also thank Tong Liu and Mohit Jain for contributions with the proteomics data. SGC is supported, in part, by the following grants: R01DK115562, U01DK10692, R01HL85757, R01DK112258, and U01OH011326. KNC is supported by NIH/NIDDK R01 grants DK103022 and DK122807. LR is supported by South Florida Veterans Affairs Foundation For Research and Education (SFVAFRE).

